# Factors impacting on the implementation of clinical management guidelines (CMGs) for high consequence infectious diseases (HCIDs) during outbreaks globally: a systematic review

**DOI:** 10.1101/2024.11.21.24317702

**Authors:** Dania Talaat Dahmash, Melina Michelen, Ishmeala Rigby, Helen Piotrowski, Robert Nartowski, Vincent Cheng, Andrew Dagens, Eli Harriss, Peter Hart, Shevin Jacobs, Keerti Gedela, Peter W Horby, Caitlin Pilbeam, Louise Sigfrid

## Abstract

**Background:** High consequence infectious disease (HCID) outbreaks are a threat to societies globally. Evidence-based clinical management guidelines (CMGs) are important tools for translating evidence into clinical practice. However, developing guidelines is resource-intensive and guidelines must remain responsive to new evidence while being accessible to clinicians. This review aims to identify factors that impact the implementation of HCID CMGs across different contexts during health emergencies.

**Methods:** A systematic review. Four databases (Ovid MEDLINE, Ovid Embase, Ovid Global Health, and Scopus) were searched until November 2021, complemented by a grey literature search conducted on November 2021. Studies that explored implementation of HCID guidelines were included, without language restriction. Two reviewers screened articles and extracted data. Data was analysed using qualitative inductive thematic analysis.

**Results:** Of 12,512 records, 28 studies were included, with most (61%, 17/28) set in high-income countries. Three overarching themes impacting HCID CMG implementation were identified: 1) Development and characteristics of CMGs, 2) Organisational and logistical factors, and 3) Realities of Implementing guidelines. Key recommendations included engaging all relevant representatives in CMG development, including those in endemic countries; integrating mechanisms for regular updates; supporting implementation by ensuring access to necessary resources (e.g., equipment, pharmaceuticals), and training; and enabling intra- and inter-organisational collaboration and communication channels. Importantly, recognising the challenges faced by staff in implementing new guidance is crucial, as is understanding the impact of a supportive environment on the effective implementation of care during emergencies.

**Conclusion:** These findings highlight the need to bridge the gap between HCID CMGs development and their real-world implementation amid health emergencies. The complex factors impacting effective implementation should be addressed beginning at the development stage, with training focused on implementation during inter-epidemic times, and ongoing implementation monitored during outbreaks. Further research to guide implementation frameworks are recommended.

**Key messages of the article:**

**What is already known on this topic:**

- Clinical management guidelines (CMGs) are important tools to guide clinical decision-making and optimise care and outcome.
- The COVID-19 pandemic showcased the need for CMGs to be rapidly responsive to new emerging evidence.
- HCID CMGs are scarce and often of low quality, and when available they frequently contain inconsistent therapeutic recommendations.
- Most CMGs are developed by high-income countries while the burden is often the largest in resource deprived settings.

**What this study adds:**

- This study highlights the gap between HCID CMGs development and their implementation in real world within emergency setting.
- The review explores the potential factors that influence the implementation process of HCID CMGs such as time, information and resource constraints.
- Key recommendations to stakeholder and CMGs developers were explored within this study such as the use of “living guidelines” to make CMGs updates more efficient, and the availability of viable alternative options for different-resourced healthcare settings to bridge the gap between the ideal situation and the local realities.
- There is a need for a clear communication and consensus on HCP expectations and obligations during health emergencies within CMGs as well as the practicalities of delivering training during emergencies need to be addressed within CMG development and implementation.

**How this study might affect research, practice or policy:**

- HCID implementation research should consider these factors impacting effective implementation when planning, from the development stage through ongoing monitoring.
- Further research and funds are needed to guide implementation frameworks.

## Introduction

High consequence infectious diseases (HCIDs) are acute infectious diseases which cause high mortality, are difficult to detect, and may not have effective prophylaxis or treatment available, potentially leading to outbreaks with severe outcomes(1). Furthermore, the COVID-19 pandemic, caused by a novel coronavirus, has demonstrated that if not effectively controlled, HCIDs have pandemic potential. Health emergencies resulting from HCIDs, can range in severity and scope from localised to widespread outbreaks. Clinical management guidelines (CMGs) are important tools that facilitate standardisation of evidence-based healthcare and guide clinical decision-making (2). The COVID-19 pandemic has demonstrated the importance of rapidly developing and implementing high-quality, evidence-based CMGs. Standardising evidence-based care can play a key role in benefiting patient care and outcomes, and also facilitate implementation of multisite clinical trials to identify optimal care strategies. The COVID-19 pandemic also illustrated the need for CMGs to be responsive to new emerging evidence (3, 4), inclusive of diverse at-risk populations, and their implementation should be supported by access to recommended treatments.

A series of systematic reviews have identified limitations in availability of HCID CMGs; many were of low quality and scope, and when available, at times contradictory supportive care and therapeutic recommendations (5–8). Most were developed by large organisations in high income countries (HICs), as defined by the World Bank, although the burden of HCIDs is generally largest in resource deprived settings(9). Developing CMGs is resource intensive, and this needs to be balanced against other priorities, particularly in resource deprived settings. CMGs developed by organisations in HICs may offer a more sustainable model if they are applicable to endemic regions and their implementation is supported. However, there may be contextual challenges to implementing guidelines in different settings. Despite efforts to develop and disseminate evidence-based CMGs, there is limited data evaluating and monitoring effective implementation of infectious diseases guidelines (10–13).

Previous studies have highlighted the complexity of how guidelines are put into practice, and how macro-meso-, and micro-level contextual factors i.e., at policy-, organisational- and inter-personal level may all influence which and how clinical guidelines are implemented (14, 15). Understanding how these contextually dependent factors impact implementation in different settings will help inform strategies to support uptake and effective implementation. Others have highlighted that multifaceted interventions are required to support clinical guideline implementation (16, 17). This is to our knowledge the first systematic review of factors impacting on the implementation of high-consequence infectious disease CMGs during outbreaks in different settings globally.

## Methods

This review was reported according to the Preferred Reporting Items for Systematic reviews and Meta-Analyses (PRISMA) guidelines (Supplementary file 1).

### Search strategy

We searched articles in Ovid MEDLINE, Ovid Embase, Ovid Global Health, and Scopus databases from inception until 16 November 2021 (Supplemental file 2). This was supplemented by a grey literature search using Google Scholar on 16^th^ of November 2021 with the first 500 records screened for inclusion. The reference lists of included studies were screened for any additional studies using the Citation chaser software (18).

### Eligibility criteria and screening

Studies describing factors impacting the implementation of HCID CMGs during health emergencies were included. The search included COVID-19, Crimean-Congo haemorrhagic fever, Ebola Virus Disease (EVD), Lassa fever, Middle East respiratory syndrome (MERS-CoV), severe acute respiratory syndrome (SARS), nipah, henipaviral diseases, Rift Valley fever, Marburg virus disease, Zika, “Disease X”, pandemic influenza, monkeypox (now mpox), chikungunya, severe fever with thrombocytopenia syndrome (SFTS), dengue fever and plague. Many of these are or have been identified as high priority pathogens for research and development by the World Health Organization (WHO) or identified as pathogens expanded into new regions in recent decades and with significant impact on public health (19). There was no exclusion based on study design or language.

Two reviewers screened the identified articles by title and abstract, followed by full text screening using the Rayyan systematic review software (20). Any disagreements were resolved by consensus or by involvement of a third reviewer.

### Data extraction and analysis

We developed a standard data extraction form on Microsoft Excel to capture included articles characteristics, including bibliography, study design, type of HCID, population, and implementation challenges and facilitators. Data extraction was performed by one researcher and verified by a second researcher. We conducted descriptive statistics to develop a summary of the included articles.

Then, we conducted inductive thematic analysis as described by Braun and Clarke (21). This consisted of two coders developing a codebook from a review of the extracted data and then independently categorizing the findings from the results and discussions of the identified articles using these codes. The codes were iteratively then reviewed and interpreted into themes and subthemes.

### Risk of bias assessment

Two reviewers independently assessed risk of bias of qualitative, quantitative, and mixed method studies using the Mixed Methods Appraisal Tool (MMAT) using quality rating scores adopted from Wranik et al (22, 23). Studies scoring between 0 to 2 points were rated as low quality; 3 to 4 points as moderate quality; 5 points as high quality. The Joanna Briggs Institute (JBI) checklist was used to assess opinion pieces, the tool assesses the validity and the motives of the opinions, the credibility of the source and the global context in terms of complementary views considered (24).

## Results

A total of 11,268 articles were identified through the search strategy, of which 28 met the selection criteria and were included in the analysis (Figure 1).

**Figure 1:**
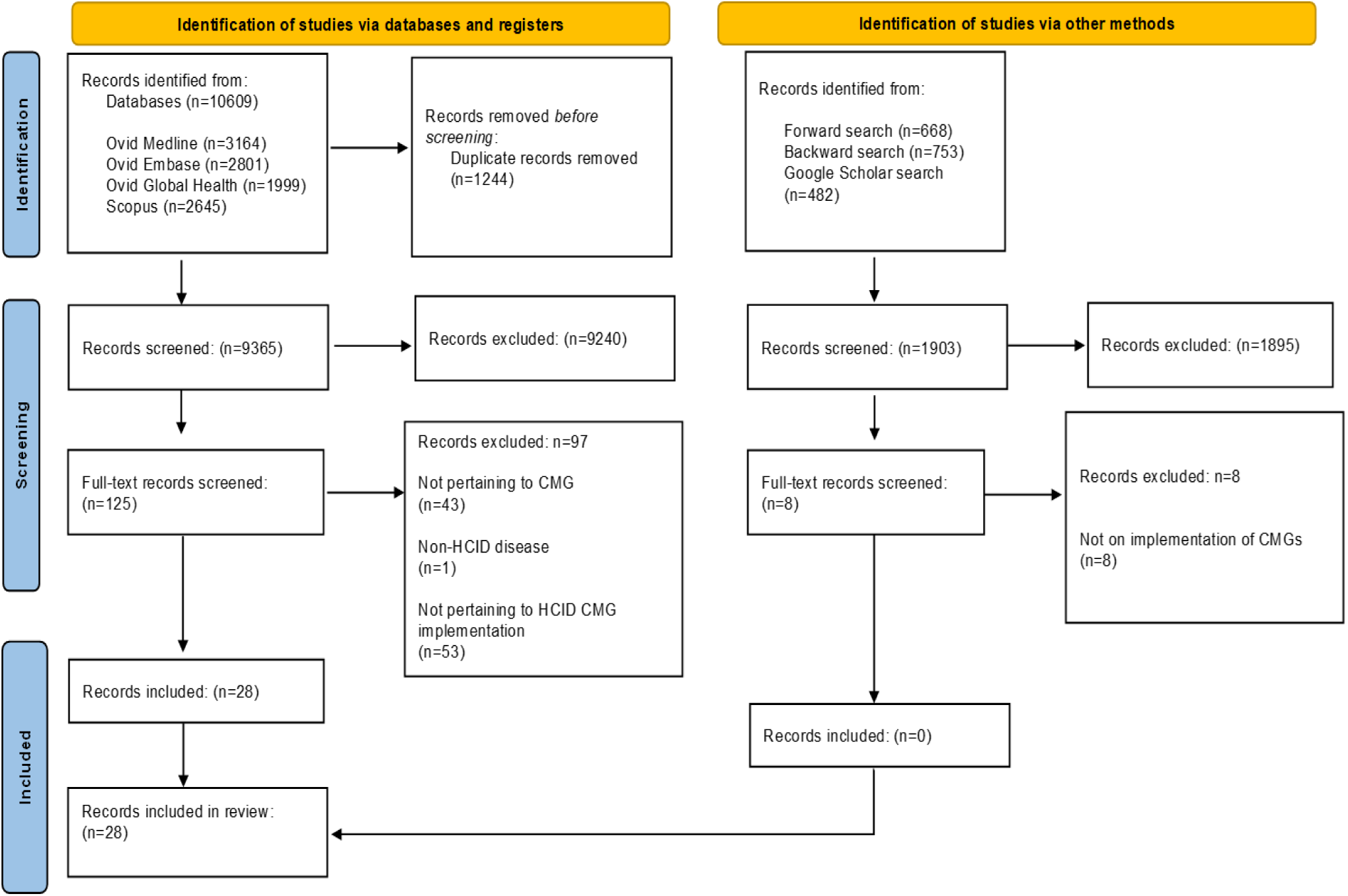
PRISMA flow chart.

### Study characteristics

Of the 28 articles included for analysis (Supplementary file 3), 12 (43%) used qualitative methods (25–36),

9 (32%) quantitative (37–45), 4 (14%) mixed methods (46–49), and 3 (11%) were opinion pieces (50–52). The majority focused on COVID-19 (29%, 8/28) or Zoonotic Influenza (25%, 7/28) and, Dengue (18%, 5/28), EVD (18%, 5/28), MERS-CoV (4%, 1/28), viral haemorrhagic fevers (4%, 1/28), and generic emerging infectious diseases (4%, 1/28). Sixty-one percent (17/28) were set in HIC (61%, 17/28) (Table 1).

**Table 1:** Type of high consequence infectious disease guideline by income setting.

| CMG focus | Total<br>N | LIC<br>n (n/N%) | MIC<br>n (n/N%) | HIC<br>n (n/N%) | Global/<br>Regional<br>n (n/N%) |
| --- | --- | --- | --- | --- | --- |
| COVID-19 | 8<br>(29%) | - | - | 7 (88) | 1 (12) |
| Dengue | 5<br>(18%) | - | 4 (80) | - | 1 (33) |
| Ebola Virus Disease (EVD) | 5<br>(18%) | 3 (60) | - | 2 (40) | - |
| Zoonotic influenza | 7<br>(25%) | - | 1 (14) | 5 (71) | 1 (14) |
| MERS-CoV | 1<br>(4%) | - | - | 1 (100) | - |
| Viral Haemorrhagic Fever (VHF) | 1<br>(4%) | - | - | 1 (100) | - |
| Emerging Infectious Diseases (EID) | 1<br>(4%) | - | - | 1 (100) | - |
| **Total n (%)** | **28** | **3 (11)** | **5 (18)** | **17 (61)** | **3 (11)** |

**Note:** CMG: Clinical Management Guidelines; COVID-19: Coronavirus disease 2019; EID: Emerging infectious diseases; EVD: Ebola Virus Disease; HIC: High Income Countries; LIC: Lower Income Countries; MIC: Middle-Income Countries; VHF: viral haemorrhagic fevers.

### Risk of bias

Of those eligible for appraisal using the MMAT tool, 60% (15/25) were rated as high quality, 24% (6/25) as moderate, and 12% (3/25) as low quality (Table 2). Most qualitative studies (92%, 11/12) were rated high quality (25–31, 33–36), while one study (8%) was rated moderate (32). Of the quantitative studies only 33.3% (3/9) were rated high quality (37, 41, 44), five studies (55.6%) were rated moderate (38–40, 43, 45) and one study (11.1%) was rated low (42). For mixed method studies, 50% (2/4) were rated high quality (46, 47) (Supplementary file 4). All studies provided a clear research question and collected data addressing the specified research questions.

**Table 2:**
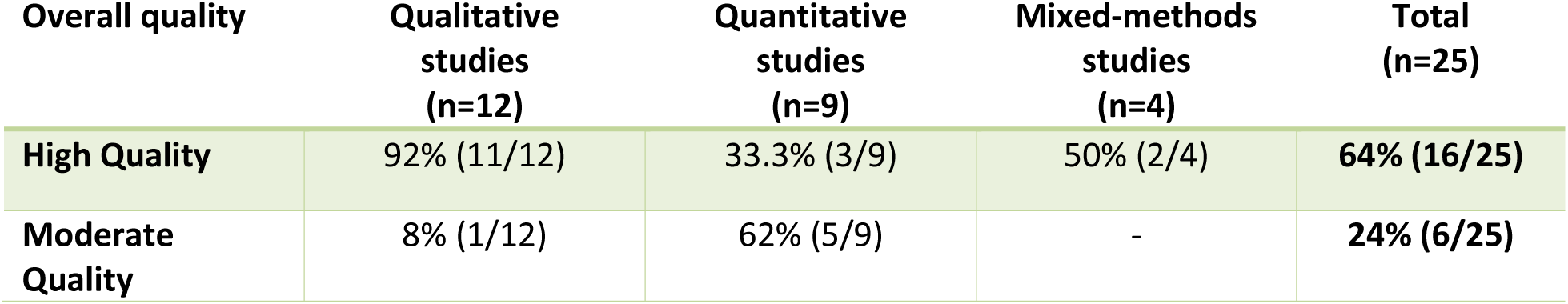
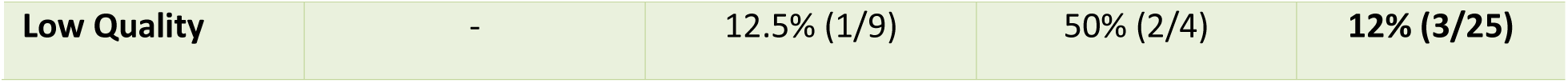
Quality of studies assesses using the MMAT tool. Twenty-five studies fit the criteria for assessment.

### Factors impacting on the implementation

Our analysis identified three overarching themes with 8 subthemes (Figure 2): 1) Development and characteristics of CMGs; 2) Organisational and logistics factors; 3) Realities of implementing guidelines.

**Figure 2:**
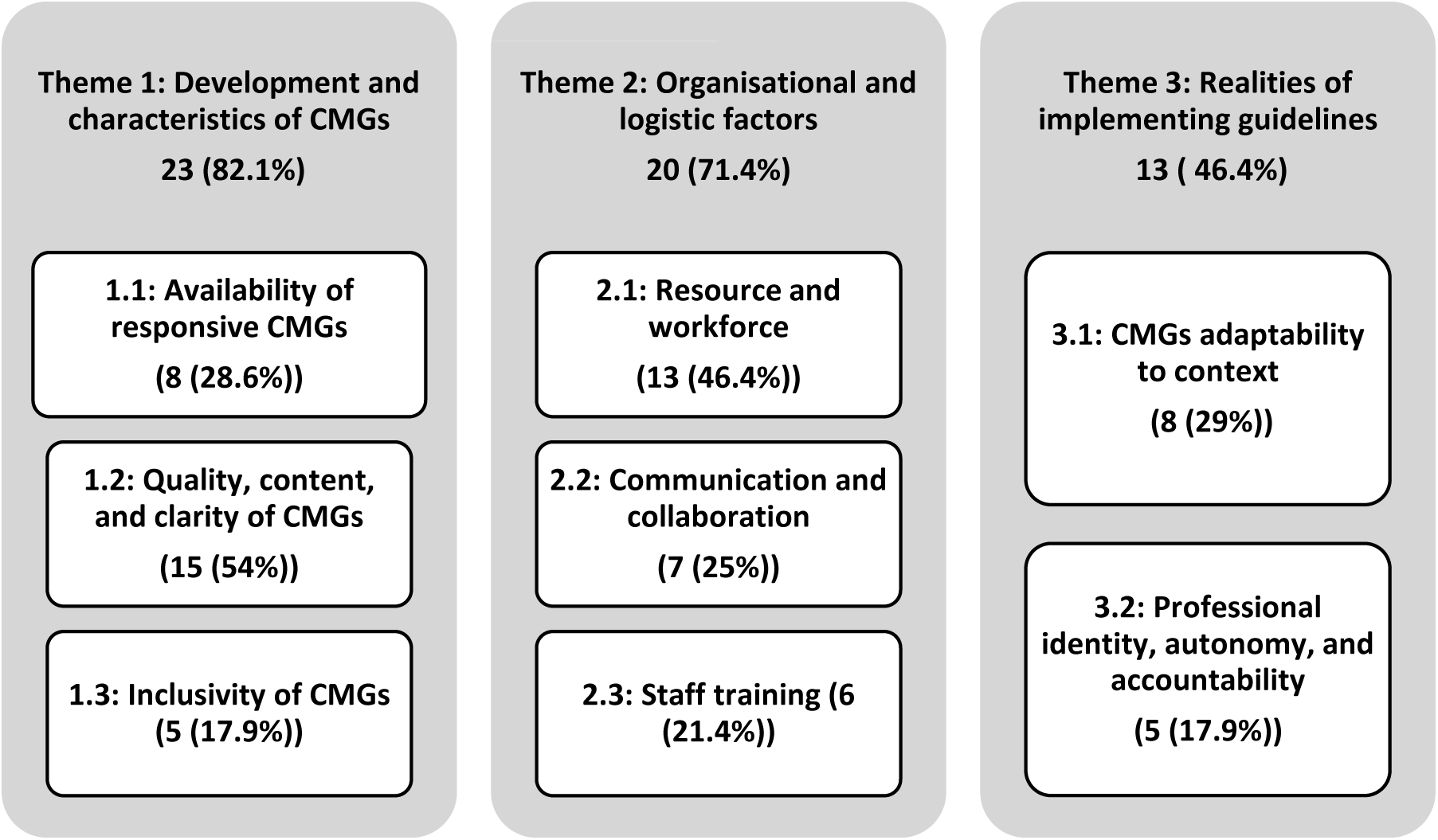
Key themes and subthemes identified. **Note: The numbers represent the numbers of articles where these were identified (n (n/N%)) where N=28 articles.**

### Theme 1: Development and characteristics of CMGs

This theme was identified in 23 (82.1%) of the articles and encompasses three sub-themes: the availability of responsive CMGs; the quality, content, and clarity in presentation; and the inclusivity of different at-risk population groups.

#### 1.1 Availability of responsive CMGs

Eight articles (28.6%) underlined the need for flexible and timely CMGs. They stressed the importance of early availability and the necessity for CMGs to be regularly updated with the best available evidence-based recommendations (25, 27, 31, 39, 44, 50–52). A study on the implementation of CMGs for influenza-like illness noted that CMGs are useful during epidemics and pandemics for facilitating timely, standardised treatment. However, despite CMGs existing, some clinicians were unaware of their availability (25). A review on lessons learnt from the H1N1 2009 pandemic in the Mediterranean region stressed that “plans should be in place for a number of scenarios well in advance” (51). The development of interim CMGs, tailored to different contexts and considering factors such as country capacity was recommended.

Four (14%) studies emphasised the commitment of resources for revisions, rapid updates as new evidence emerges, and re-dissemination to frontline clinicians at the development stage. (31, 44, 51, 52). This approach can enhance equity in access to the best available care recommendations and prevent the use of outdated information.

#### 1.2 Quality, content, and clarity of CMGs

This subtheme was present in fifteen articles (54%). Healthcare professionals (HCPs) expressed concerns about the quality of CMGs (26, 28, 29, 33, 48). Challenges included the high volume of information received during emergencies, rapidly changing guidelines, inconsistent and sometimes contradictory recommendations, limited clarity of guidelines, and delays in disseminating new evidence (26, 28, 29, 33, 48). Participants in one study emphasized that discrepancies and large amounts of rapidly received information for EVD, both locally and from organizational and government levels, resulted in the dismissal of information, as clinicians could not assimilate it all in the limited time available (26). They perceived guidelines as being developed ‘on the fly’, resulting in uncertainty, confusion, and lack of trust in the information received (26). Respondents in a survey on pandemic influenza management reported that guideline content was inconsistent between different pandemic management levels and sometimes contradictory, contributing to confusion amongst clinicians (29). Participants in a study on H1N1 reported “there was too much information and too many sources” (48). Similarly, midwives caring for pregnant women with EVD, expressed being ‘bombarded’ with opinions and information from people without obstetrical competency (28).

Ten studies (36%) emphasized the need for CMGs to provide clear recommendations (25, 27, 30, 33–35, 39, 42, 49, 51). Studies discussed the content and user-friendliness of CMGs as important for effective implementation (25, 27, 30, 33–35, 42, 44, 49). A study of CMGs for COVID-19 in Qatar demonstrated that the clarity of CMGs is strongly associated with overall primary care physician job satisfaction (44). Another study reported that CMGs were too complex and unwieldy, leading to HCPs disregarding them (33, 49). One study emphasized the importance of CMGs including elements such as treatment and referral criteria, infection control policies, and guidance on required infrastructure, capacity, and skills for policymakers to enhance preparedness for epidemics and pandemics (25).

HCPs perceiving CMGs as credible and trustworthy was highlighted as integral for implementation, underlining that CMGs need to back up recommendations with scientific evidence to be perceived as trustworthy (27, 30, 33, 42). Additionally, it was mentioned that CMGs need to be developed by “respected, trusted sources.” (27, 30). However, clinicians cited a lack of robust evidence as a barrier to implementing CMGs in practice, an issue at-times even for CMGs produced by reputable organization such as the World Health Organisation and US Center for Disease Control (33).

#### 1.3 Inclusivity of CMGs

The lack of inclusivity regarding specific population groups, such as children, elderly, and people with comorbidities was highlighted as a challenge for implementation in five articles (17.9%) (27, 28, 32, 34, 50). For instance, when implementing EVD guidelines, HCPs reported frustration over the lack of clear guidelines for caring for pregnant women with EVD, leading to practical and ethical dilemmas, prompting them to seek guidance from alternative sources (28). Another study recommended clearer delineation of target patient groups based on demographics and specific risk factors (e.g., ventilated patients) (27). One study emphasised the importance of including guidance for marginalized groups, such as indigenous populations, recognising that existing healthcare inequities and other disparities may already place them at a higher risk of severe outcomes from HCIDs (50). Even guidelines aimed at specific groups, like pregnant women with COVID-19, were found by medical staff to be of limited scope and not comprehensive enough to cover all aspects of care delivery (32).

### Theme 2: Organisational and logistic factors

This theme, identified in 20 (71.4%) of the included articles, encompasses three subthemes describing resource and workforce availability, communication and collaboration, and HCP training.

#### 2.1 Resource and workforce

Thirteen articles (46.4%) identified workforce capacity and insufficient material and other resources, including time, therapeutics, protective equipment, laboratory facilities, and access to electronic health records, as key factors influencing CMG implementation (25, 27, 29, 32, 37, 39–41, 45–47, 49, 50). In one study, 31% of HCPs reported inadequate access to the resources required for the diagnosis and treatment of Dengue fever (41). These limitations were especially problematic during emergency situations, where time constraints added extra pressure (27). Another study highlighted the importance of capacity planning for pandemics, since resource reallocation might have a negative impact on other essential services. This was a particular concern for rural communities where public health capacity may be limited (50).

#### 2.2 Communication and collaboration

The impact of communication and organisational collaboration was highlighted in seven articles (25%) (27, 29, 34, 36, 39, 40, 47).

Insufficient intra- and inter organisational communication was cited as a challenge for effective guideline implementation (34, 39, 47). A key recommendation for effective CMG implementation was to enhance communication and collaboration between different healthcare representatives (34, 35, 37, 38, 46, 48, 50, 51). Further, two studies recommended greater centralisation and improved utilisation of new technologies and the internet for streamlined, effective communication routes between those on frontline workers and authorities (35, 48).

A study on COVID-19 in Saudi Arabia identified strong leadership and organisational support as additional facilitators (37). One study emphasised the need for strong unit-based support for evidence-based practice to facilitate the implementation of a new guideline during a pandemic, as “staff engagement was strengthened when unit-level leadership and education were supported by a value for EBP” (40).

#### 2.3 Staff training

Six articles (21.4%%) underscored a lack of training as a major challenge faced by HCPs when implementing CMGs during emergencies (37, 41, 43, 46, 47). Recommended training varied from informational (e.g. guideline content) to practical (e.g. outbreak drills) to improve awareness of the guideline’s content and practice implementation to identify and address challenges before outbreaks occur. For instance, in a study with participants caring for patients with VHF, insufficient personal experience led to uncertainty and anxiety, highlighting a need for frequent staff training to facilitate CMG implementation (47). Another study focused on Dengue guidelines, showed that despite high levels of knowledge regarding symptoms and treatment, HCPS still needed additional training in prevention, diagnosis, and admission criteria, and on the WHO Dengue Guidelines (41). A quality improvement intervention study showed that training on Dengue guidelines during the off-peak season increased post-intervention compliance on all CMG components, resulting in decreased mortality and length of hospital stay during peak season (38). Educational interventions were found not only to improve patient outcomes, but also to guide appropriate patient management and resource utilisation, such as proper use of platelet transfusions and antibiotics (38). Clinicians in another study recommended they “practice using the protocol during drills to make sure the details of the protocol would work”(27).

### Theme 3: Realities of implementing guidelines

The realities of implementing CMGs reflect limitations in adapting recommendations into contextual practice, with factors relating to HCPs professional identity described in 13 (46.4%) studies.

#### 3.1 Adaptability to context

Eight (29%) articles underscored the importance of adapting CMGs to fit local contexts (29, 31, 35, 40, 42, 48, 50, 51). There was a recognised need to better balance theory and clinical practice, with one study noting CMGs are “no more than a paper exercise without considering possible obstacles in real world situations.” (31). During the 2009 H1N1 influenza pandemic, physicians reported difficulties reconciling public health guidance with the realities of their practice, with one respondent noted “the biggest challenge was not our internal resource but really trying to figure out how we would apply the recommendations to our practice” (29). Similarly, another study identified implementation challenges stemming from guidelines perceived as too rigid for regional and local context (48). Additional challenges identified were discrepancies between international guidelines and their practical implementation at the local-level (35).

While consistent messaging from various authorities was seen as beneficial, there is a call for flexibility to adapt guidelines appropriately.(50) To address delays and challenges in guideline adoption, one study recommended adapting international guidelines to national and local context (35). Another study recommended involving representatives from endemic countries in the CMG development process to facilitate applicability and relevance (51). Engaging planning committees at various levels, such as local and provincial, in routine reviews of the guideline implementation process between outbreaks was also suggested to identify gaps and address implementation bottlenecks. Even at the local level, one survey highlighted differences in opinions and practices among clinicians; to mitigate this, they proposed that guidelines should “suit the local context, based on epidemiological evidence, clinical features, and management” (42). Further, WHO guidelines should consider local context applicability, by engaging local representatives, including experienced clinicians to share evidence and experiences to inform CMG development and implementation support (42). Similarly, one study suggests that understanding the different unit-based attitudes and beliefs towards adoption of new practices was also important (40).

#### 3.2 Professional identity, autonomy, and accountability

Five articles (17.9%) discussed the role of HCPs’ autonomy and wellbeing for effective CMG implementation, particularly in the context of the burden imposed by health emergencies (27, 28, 32, 34, 47). One study noted staff experiences physical and psychological challenges during pandemics, including fatigue, stress, uncertainty, and worry (32). Another study addressed concerns about exposure risk, suggesting accommodations for staff facing heightened exposure risk (47).

Three articles highlighted the complexity and tensions associated with implementing HCID CMGs (27, 28, 34). Using CMGs effectively demands significant clinical expertise, yet adherence to CMGs may also be perceived as undermining expertise. Junior clinicians noted “adherence to guidelines can sit uneasily, with the need to demonstrate ones’ own professional competence” (34). Another study noted that guidelines could potentially threaten HCPs’ “sense of self in the professional sphere”, with HCPs noting “…you’re no longer a physician at this point, you are just following orders” (27). Similarly, midwives in another study described relying on their own “competency and professional creativity”, when CMGs were considered too rigid and restrictive (28).

### Summary of recommendations

This literature review has identified areas for improvement and key recommendations for future CMG development and implementation. These are summarised in Table 3.

**Table 3:** Summary of key recommendations identified.

| Key areas of improvements | Key recommendations identified |
| --- | --- |
| **CMG quality, content, and clarity** | - Produce clear, adaptable, and inclusive CMGs.<br>- Include evidence-based, up-to-date treatment recommendations and referral criteria for clinicians.<br>- Include information (e.g., infection control policies, guidance on required infrastructure, capacity, and skills) targeted at policy makers to improve preparedness during emergencies.<br>- Engage representatives with experience of managing HCIDs in different context to facilitate applicability and implementation. |
| **Communication and collaboration** | - Establish strong internal and external collaboration and communication channels in preparedness time.<br>- Prioritise internet and other communication systems and pathways to share information rapidly with all stakeholders. |
| **Training** | - Conduct staff training on CMGs prior to an emergency occurring/out of peak epidemic seasons, with regular updates.<br>- Conduct practice implementation exercises to identify bottlenecks to address in inter-epidemic times. |
| **Resources and leadership** | - Enable strong leadership for resource and workforce management and to provide general support to HCPs during emergencies.<br>- Review resources required for implementation of CMGs.<br>- Ensure evidence-based CMGs are available to frontline clinicians across health centers.<br>- Foster a work culture that supports use of evidence-based CMGs to inform practice and equity in access to evidence-based recommendations.<br>- Facilitate good collaboration and communications internally and across organisations. |
| **Adaptability to context** | - Involve diverse representatives, including health workers, in identifying local challenges to address prior to outbreaks.<br>- Support cross-country development and adaptation of CMGs.<br>- Ensure context-specific factors are addressed and representatives from endemic countries are included in the development and dissemination. |

## Discussion

There is limited published data on factors affecting HCID CMG development and implementation during health emergencies. To address this gap, this review explores potential factors that influence the implementation process of HCID CMGs. Our findings align with similar findings on CMGs conducted outside of emergency settings (53). Whilst the factors described were similar across outbreaks and settings, additional challenges arise with emerging pathogens and in lower-resourced settings, due to time, information, and resource constraints. Reflecting on the articles and their recommendations, we present further considerations for CMG development and implementation.

### Establish mechanisms for CMGs to be updated and disseminated

Updating CMGs is a resource-intensive task. Therefore, approaches to make updates more efficient should be considered, including use of ‘living’ guidelines (54). The findings acknowledge that regular updates of CMGs are crucial for effective implementation, given the rapid emergence of new evidence. Furthermore, dissemination can be challenging in some settings, such as where HCPs may have limited internet access, thus limiting the timely accessibility of obtaining up-to-date CMGs (55). Diversifying dissemination strategies to include both on- and off-line alternatives, whilst also ensuring content consistency and avoiding overwhelming HCPs with excessive information, can help address this issue.

### Ensure CMGs are adapted to context and identify early potential bottlenecks to implementation

The adaptability of CMGs to local contexts is another key factor affecting the implementation process (56, 57). A CMG that is well-integrated in one setting may not be as well-integrated in another (58). Hence, involving diverse guideline developers from different contexts is important for optimising future implementation. One study identified that narrow skill representation in the development team could lead to lower CMG quality and uptake by clinicians (59). Often, HCPs from low-income settings are not included on guideline development boards or in the piloting of CMGs, resulting in guidelines that are difficult or unrealistic to implement across diverse settings. This highlights the need for CMGs to be tailored to fit local epidemiological and resource context (60, 61).

Additionally, implementation is also hindered by shortages of facilities and material resources required by the CMGs (15, 62, 63). A systematic review of factors influencing CMG implementation suggests that CMGs which do not require specific resources have a greater chance of implementation (57). This is particularly the case for HCIDs and during emergencies, where resource shortages are likely. CMGs should therefore include viable alternatives and ‘next best’ options for differently-resourced health care settings, and to acknowledge the gap between ideal situations and shifting local realities.

### Support health care professional wellbeing during emergencies

Aligning with our findings, previous literature indicates that inconsistent and difficult-to-follow CMGs may burden HCPs, thus hindering implementation(64, 65) A study on the stress drivers among HCPs during COVID-19 identified changing CMGs, resource shortages, and fear of transmission as key factors (15). Similarly, our findings show that HCPs expressed concerns about exposure, liability, and stress during health emergencies. Therefore, clear communication and consensus on HCP expectations and obligations during health emergencies within CMGs are essential, addressing both legal and ethical considerations (66). Additionally, providing opportunities for collective debriefing post-emergency can support staff in processing events and accessing necessary support.

Our findings highlighted that HCPs expressed concerns about their professional identity and autonomy; perceiving CMGs to challenge this (18, 67–71). Effective implementation should promote “guideline adoption as consistent with their professional autonomy” (72). This can be facilitated through organisational support, strong leadership, and cultivating a supportive working culture and peer groups, all recognised as crucial for the implementation and sustainability of CMGs (56, 57).

### Provide on-going and sustainable training for HCPs on CMGs

HCP training was frequently addressed in CMG implementation studies (15, 73, 74). The practicalities of delivering training during emergencies and in different settings (e.g., high- and low-income) need to be addressed within CMG development and implementation processes. During health emergencies, HCPs can be redeployed and required to perform new tasks with limited training time; as such, training strategies should focus on maximising transferable skills (60, 63, 75–77). Lastly, early preparedness training is essential to effectively respond during an emergency. Hence, mock drills may be useful, also to enhance confidence in and understandings of putting guidelines into practice, and identify potential limitations in doing so (78). Finally, There is a need for collaborative development of clinical trials and CMGs with endemic countries from the outset to facilitate implementation of well-designed trials with capacity to address local needs and influence local policies during outbreaks.

### Strengths and limitations

This review is not without limitations. Firstly, although the search strategy was trialled before being finalised, it is still possible to have missed important literature, considering some endemic HCIDs in countries which may not have published data on CMG implementation using conventional methods (i.e., unpublished or in internal government documentation). Second, whilst there were no language restrictions, the literature search was conducted in English, potentially leading to the retrieval of articles only in English. Finally, articles were not excluded based on their quality rating. However, a key strength of this study is its global perspective, addressing the factors that affect the implementation of CMGs during HCID emergencies. This is the first systematic review to describe HCID CMG implementation factors during emergencies and it includes data from 18 different countries. Our review adheres to rigorous methods, including a systematic search strategy unrestricted by date or language and explicit inclusion criteria. Therefore, the findings present the most comprehensive and internationally relevant data on HCID CMG implementation during emergencies worldwide.

## Conclusion

Health emergencies are high-pressure, high-stakes contexts in which effective CMG implementation is crucial, as CMGs help facilitate the provision of evidence-based healthcare, particularly useful when evidence is limited. The gap between written guidelines, and the (local) lived practice of implementing guidelines during a health emergency must be acknowledged and addressed. Factors affecting implementation of CMGs for HCIDs are multifaceted and can act as either barriers or facilitators, depending on the context. These factors can inform and be integrated into policy and practice when designing, developing, and implementing guidelines, to ensure effective implementation and uptake of CMGs in different contexts. To support the production of context-appropriate, acceptable CMGs and their uptake, we therefore recommend that CMG development at all levels involve diverse representatives and focus on their implementation in differently resourced settings. Research on interventional studies should address the challenges and determine the effectiveness of interventions targeting the enabling factors.

## Declarations

**Ethics approval and consent to participate.**

Not applicable

## Consent for publication

Not applicable

## Availability of data and materials

All data generated or analysed during this study are included in this published article and its supplementary information files.

## Authors contributions

LS, KG, CP, DD, MM and IR informed the study protocol. DD, MM, and LS led on writing the manuscript with input from all co-authors. EH and VC carried out the database search with input from DD, MM, and IR. HP, RN, DD, MM, IR, EC and SK conducted the grey literature search. RN, DD, IR, and MM screened the retrieved articles for inclusion. RN, DD, IR, MM extracted the data and completed the risk of bias analysis. LS, DD, CP, MM, IR lead on the data analysis and presentation of the results. LS, DD, MM, IR, CP, informed the interpretation of the results. LS, PWH, KG and STJ provided overall supervision, leadership and advice. PWH, HG, STJ, TF, PH, LS, AD and LB conceptualised the project. All authors reviewed and approved the final version of the manuscript. The first authors (DD, MM) are the guarantors who had access to all the data, and accept responsibility for the work, the conduct of the study and the decision to publish.

## Data Availability

All data produced in the present study are available upon reasonable request to the authors.

## Acknowledgment

Thanks to the ISARIC Global Support Centre for logistical and administrative support and all the members of the ISARIC Global Clinical Research Networks who completed the survey.

## References

1. GOV.UK. High consequence infectious diseases (HCID) 2024 [

2. Steinberg E, Greenfield S, Wolman DM, Mancher M, Graham R. Clinical practice guidelines we can trust: national academies press; 2011.

3. Siemieniuk R, Rochwerg B, Agoritsas T, Lamontagne F, Leo Y-S, Macdonald H, et al. A living WHO guideline on drugs for covid-19. BMJ: British Medical Journal. 2020;370:1–14.

4. Nicola M, O’Neill N, Sohrabi C, Khan M, Agha M, Agha R. Evidence based management guideline for the COVID-19 pandemic-Review article. International Journal of Surgery. 2020;77:206–16.

5. Webb E, Rigby I, Michelen M, Dagens A, Cheng V, Rojek AM, et al. Availability, scope and quality of monkeypox clinical management guidelines globally: a systematic review. BMJ global health. 2022;7(8):e009838.

6. Webb E, Michelen M, Rigby I, Dagens A, Dahmash D, Cheng V, et al. An evaluation of global Chikungunya clinical management guidelines: a systematic review. EClinicalMedicine. 2022;54.

7. Rigby I, Michelen M, Dagens A, Cheng V, Dahmash D, Harriss E, et al. Standard of care for viral haemorrhagic fevers (VHFs): a systematic review of clinical management guidelines for high-priority VHFs. The Lancet Infectious Diseases. 2023.

8. Rigby I, Michelen M, Cheng V, Dagens A, Dahmash D, Lipworth S, et al. Preparing for pandemics: a systematic review of pandemic influenza clinical management guidelines. BMC medicine. 2022;20(1):1–16.

9. Bank TW. The World by Income and Region 2023 [Available from: https://datatopics.worldbank.org/world-development-indicators/the-world-by-income-and-region.html.

10. Zhao J, Varin MD, Graham ID. Guidelines do not self-implement: time for a research paradigm shift from massive creation to effective implementation in evidence-based medicine research in China. BMJ Evidence-Based Medicine. 2020;25(4):118–9.

11. Feder G, Eccles M, Grol R, Griffiths C, Grimshaw J. Using clinical guidelines. Bmj. 1999;318(7185):728-30.

12. Fretheim A, Schünemann HJ, Oxman AD. Improving the use of research evidence in guideline development: 15. Disseminating and implementing guidelines. Health Research Policy and Systems. 2006;4(1):1–4.

13. Jacob ST, Lim M, Banura P, Bhagwanjee S, Bion J, Cheng AC, et al. Integrating sepsis management recommendations into clinical care guidelines for district hospitals in resource-limited settings: the necessity to augment new guidelines with future research. BMC medicine. 2013;11:1–7.

14. Correa VC, Lugo-Agudelo LH, Aguirre-Acevedo DC, Contreras JAP, Borrero AMP, Patiño-Lugo DF, Valencia DAC. Individual, health system, and contextual barriers and facilitators for the implementation of clinical practice guidelines: a systematic metareview. Health Research Policy and Systems. 2020;18(1):74.

15. Fischer F, Lange K, Klose K, Greiner W, Kraemer A. Barriers and Strategies in Guideline Implementation—A Scoping Review. Healthcare. 2016;4(3):36.

16. Wensing M, Grol R. Knowledge translation in health: how implementation science could contribute more. BMC Medicine. 2019;17(1):88.

17. Rapport F, Clay-Williams R, Churruca K, Shih P, Hogden A, Braithwaite J. The struggle of translating science into action: Foundational concepts of implementation science. Journal of Evaluation in Clinical Practice. 2018;24(1):117–26.

18. Haddaway N, Grainger M, Gray C. citationchaser: an R package and Shiny app for forward and backward citations chasing in academic searching. Zenodo. 2021.

19. WHO. Prioritizing diseases for research and development in emergency contexts. 2020.

20. Ouzzani M, Hammady H, Fedorowicz Z, Elmagarmid A. Rayyan—a web and mobile app for systematic reviews. Systematic Reviews. 2016;5(1):210.

21. Clarke V, Braun V. Successful qualitative research: A practical guide for beginners. Successful qualitative research. 2013:1–400.

22. Hong QN, Fàbregues S, Bartlett G, Boardman F, Cargo M, Dagenais P, et al. The Mixed Methods Appraisal Tool (MMAT) version 2018 for information professionals and researchers. Education for Information. 2018;34:1–7.

23. Wranik WD, Hayden JA, Price S, Parker RM, Haydt SM, Edwards JM, et al. How best to structure interdisciplinary primary care teams: the study protocol for a systematic review with narrative framework synthesis. Systematic Reviews. 2016;5:1–7.

24. Moola S, Munn Z, Tufanaru C, Aromataris E, Sears K, Sfetcu R, et al. Chapter 7: Systematic reviews of etiology and risk. Joanna briggs institute reviewer’s manual The Joanna Briggs Institute. 2017;5:217–69.

25. Ahankari AS, Myles PR, Tsang S, Khan F, Atre S, Langley T, et al. A qualitative study exploring factors influencing clinical decision-making for influenza-like illness in Solapur city, Maharashtra, India. Anthropology & medicine. 2019;26(1):65–86.

26. Broom J, Broom A, Bowden V. Ebola outbreak preparedness planning: a qualitative study of clinicians’ experiences. public health. 2017;143:103–8.

27. Chuang E, Cuartas PA, Powell T, Gong MN. “We’re not ready, but i don’t think you’re ever ready.” Clinician perspectives on implementation of crisis standards of care. AJOB empirical bioethics. 2020;11(3):148–59.

28. Erland E, Dahl B. Midwives’ experiences of caring for pregnant women admitted to Ebola centres in Sierra Leone. Midwifery. 2017;55:23–8.

29. Filice CE, Vaca FE, Curry L, Platis S, Lurie N, Bogucki S. Pandemic planning and response in academic pediatric emergency departments during the 2009 H1N1 influenza pandemic. Academic Emergency Medicine. 2013;20(1):54–62.

30. Kurotschka PK, Serafini A, Demontis M, Serafini A, Mereu A, Moro MF, et al. General practitioners’ experiences during the first phase of the COVID-19 pandemic in Italy: a critical incident technique study. Frontiers in Public Health. 2021;9:623904.

31. Lam SK, Kwong EW, Hung MS, Pang SM. Bridging the gap between guidelines and practice in the management of emerging infectious diseases: A qualitative study of emergency nurses. Journal of clinical nursing. 2016;25(19-20):2895–905.

32. González A, Pinto PH, Maldonado S, Villalobos I, Sierra N, Melgosa I. Analysis of the care management protocol for COVID pregnant women and detection of improvement proposals applying clinical simulation methodology. Revista Española de Anestesiología y Reanimación (English Edition). 2020;67(9):487–95.

33. Phipps E, Watson C, Mearkle R, Lock S. Influenza in carehome residents: applying a conceptual framework to describe barriers to the implementation of guidance on treatment and prophylaxis. Journal of Public Health. 2020;42(3):602–9.

34. Wharton-Smith A, Green J, Loh EC, Gorrie A, Omar SFS, Bacchus L, Lum LCS. Using clinical practice guidelines to manage dengue: a qualitative study in a Malaysian hospital. BMC infectious diseases. 2019;19(1):1–10.

35. Gesser-Edelsburg A, Mordini E, James JJ, Greco D, Green MS. Risk communication recommendations and implementation during emerging infectious diseases: a case study of the 2009 H1N1 influenza pandemic. Disaster medicine and public health preparedness. 2014;8(2):158–69.

36. Loignon C, Nouvet E, Couturier F, Benhadj L, Adhikari NK, Murthy S, et al. Barriers to supportive care during the Ebola virus disease outbreak in West Africa: Results of a qualitative study. PLoS One. 2018;13(9):e0201091.

37. Alanezi F. Factors affecting the adoption of e-health system in the Kingdom of Saudi Arabia. International Health. 2021;13(5):456–70.

38. Balkrishnan A, Panda PK, RMPandey AB, Aggarwal P, Vikram NK, Dar L, Wig N. Compliance of WHO Guideline on Dengue Management among Indian Patients: An Interventional Quality Improvement Study. Journal of The Association of Physicians of India. 2019;67:30.

39. Battista M-C, Loignon C, Benhadj L, Nouvet E, Murthy S, Fowler R, et al. Priorities, barriers, and facilitators towards international guidelines for the delivery of supportive clinical care during an Ebola outbreak: a cross-sectional survey. Viruses. 2019;11(2):194.

40. Francisco MA, Pierce NL, Ely E, Cerasale MT, Anderson D, Pavkovich D, et al. Implementing prone positioning for COVID-19 patients outside the intensive care unit. Journal of Nursing Care Quality. 2021;36(2):105–11.

41. Handel AS, Ayala EB, Borbor-Cordova MJ, Fessler AG, Finkelstein JL, Espinoza RXR, et al. Knowledge, attitudes, and practices regarding dengue infection among public sector healthcare providers in Machala, Ecuador. Tropical diseases, travel medicine and vaccines. 2016;2(1):1–10.

42. Kularatne S. Survey on the management of dengue infection in Sri Lanka: opinions of physicians and pediatricians. Southeast Asian journal of tropical medicine and public health. 2005;36(5):1198.

43. Alqahtani JS, Mendes RG, Aldhahir A, Rowley D, AlAhmari MD, Ntoumenopoulos G, et al. Global current practices of ventilatory support management in COVID-19 patients: an international survey. Journal of multidisciplinary healthcare. 2020:1635–48.

44. Ismail M, Joudeh A, Neshnash M, Metwally N, Seif MH, Al Nuaimi A, et al. Primary health care physicians’ perspective on COVID-19 pandemic management in Qatar: a web-based survey. BMJ open. 2021;11(9):e049456.

45. Driver JA, Strymish J, Clement S, Hayes B, Craig K, Cervera A, et al. Front-Line innovation: Rapid implementation of a nurse-driven protocol for care of outpatients with COVID-19. Journal of Clinical Nursing. 2021;30(11-12):1564–72.

46. Barniol J, Gaczkowski R, Barbato EV, da Cunha RV, Salgado D, Martínez E, et al. Usefulness and applicability of the revised dengue case classification by disease: multi-centre study in 18 countries. BMC infectious diseases. 2011;11:1-12.

47. Fryk JJ, Tong S, Marshall C, Rajkhowa A, Buising K, MacIsaac C, et al. Knowledge, attitudes and practices of healthcare workers within an Australian tertiary hospital to managing high-consequence infectious diseases. Infection, disease & health. 2021;26(2):95–103.

48. Nhan C, Laprise R, Douville-Fradet M, Macdonald ME, Quach C. Coordination and resource-related difficulties encountered by Quebec’s public health specialists and infectious diseases/medical microbiologists in the management of A (H1N1)-a mixed-method, exploratory survey. BMC Public Health. 2012;12(1):1–8.

49. Raffetin A, Ortmans C, Worms B, Cadwallader J-S. French guidelines for the outpatient management of Ebola virus disease: Applicability by family physicians. Médecine et Maladies Infectieuses. 2018;48(8):526–32.

50. Bandara T, Musto R, Kancir J, Neudorf C, editors. Public health physician perspectives on developing and deploying clinical practice guidelines during the 2009 H1N1 pandemic. Healthcare Management Forum; 2020: SAGE Publications Sage CA: Los Angeles, CA.

51. Haq Z, Malik M, Khan W. The H1N1 influenza pandemic of 2009 in the Eastern Mediterranean Region: lessons learnt and future strategy. EMHJ-Eastern Mediterranean Health Journal. 2016;22(7):548–25.

52. Omrani A, Shalhoub S. Middle East respiratory syndrome coronavirus (MERS-CoV): what lessons can we learn? Journal of hospital infection. 2015;91(3):188–96.

53. Sarrafzadegan N, Shahidi S, Kholenjani FB. The Challenges of Developing Clinical Practice Guidelines. Iranian Journal of Nursing and Midwifery Research. 2023;28(5):631.

54. Panteli D, Legido-Quigley H, Reichebner C, Ollenschläger G, Schäfer C, Busse R. Clinical practice guidelines as a quality strategy. Improving healthcare quality in Europe. 2019:233.

55. Gautham M, Iyengar MS, Johnson CW. Mobile phone–based clinical guidance for rural health providers in India. Health informatics journal. 2015;21(4):253–66.

56. Marchionni C, Ritchie J. Organizational factors that support the implementation of a nursing best practice guideline. Journal of nursing management. 2008;16(3):266–74.

57. Francke AL, Smit MC, de Veer AJ, Mistiaen P. Factors influencing the implementation of clinical guidelines for health care professionals: a systematic meta-review. BMC medical informatics and decision making. 2008;8(1):1–11.

58. Wallisch JS, Pang D, Carcillo JA, Aneja RK. Implementation of guidelines to treat pediatric sepsis: Cookbook medicine or the force awakens! Pediatric Critical Care Medicine. 2016;17(9):884–5.

59. Guo Y, Adelstein BA, Rubin GL. Availability and development of guidelines in a tertiary teaching hospital. Journal of Evaluation in Clinical Practice. 2007;13(4):632–8.

60. Maaløe N, Ørtved AMR, Sørensen JB, Dmello BS, van den Akker T, Kujabi ML, et al. The injustice of unfit clinical practice guidelines in low-resource realities. The Lancet Global Health. 2021;9(6):e875–e9.

61. Norris SL, Ford N. Improving the quality of WHO guidelines over the last decade: progress and challenges. The Lancet Global Health. 2017;5(9):e855–e6.

62. Otgon B, Lundeg G, Tsenddorj G, Jochberger S, Grander W, Baelani I, et al. Nationwide survey on resource availability for implementing current sepsis guidelines in Mongolia. Bulletin of the World Health Organization. 2010;88:839–46.

63. Kredo T, Cooper S, Abrams A, Muller J, Volmink J, Atkins S. Using the behavior change wheel to identify barriers to and potential solutions for primary care clinical guideline use in four provinces in South Africa. BMC health services research. 2018;18(1):1–12.

64. Jun J, Kovner CT, Stimpfel AW. Barriers and facilitators of nurses’ use of clinical practice guidelines: an integrative review. International journal of nursing studies. 2016;60:54–68.

65. Morris AH. Developing and implementing computerized protocols for standardization of clinical decisions. Annals of internal medicine. 2000;132(5):373–83.

66. Johnson SB, Butcher F. Doctors during the COVID-19 pandemic: what are their duties and what is owed to them? Journal of Medical Ethics. 2021;47(1):12–5.

67. Timmermans S. From autonomy to accountability: the role of clinical practice guidelines in professional power. Perspectives in biology and medicine. 2005;48(4):490–501.

68. Lau R, Stevenson F, Ong BN, Dziedzic K, Treweek S, Eldridge S, et al. Achieving change in primary care—causes of the evidence to practice gap: systematic reviews of reviews. Implementation Science. 2015;11(1):1–39.

69. Baradaran-Seyed Z, Nedjat S, Yazdizadeh B, Nedjat S, Majdzadeh R. Barriers of clinical practice guidelines development and implementation in developing countries: a case study in Iran. International journal of preventive medicine. 2013;4(3):340.

70. Lin KY. Physicians’ perceptions of autonomy across practice types: is autonomy in solo practice a myth? Social Science & Medicine. 2014;100:21–9.

71. Klazinga N. Compliance with practice guidelines: clinical autonomy revisited. Health Policy. 1994;28(1):51–66.

72. Adler PS, Kwon SW. The mutation of professionalism as a contested diffusion process: clinical guidelines as carriers of institutional change in medicine. Journal of Management Studies. 2013;50(5):930–62.

73. Gagliardi AR, Alhabib S, Group motGINIW. Trends in guideline implementation: a scoping systematic review. Implementation Science. 2015;10:1–11.

74. Yazdani M, Sabetian G, Ra’ofi S, Roudgari A, Feizi M. A comparative study of teaching clinical guideline for prevention of ventilator-associated pneumonia in two ways: face-to-face and workshop training on the knowledge and practice of nurses in the Intensive Care Unit. Journal of Advances in Medical Education & Professionalism. 2015;3(2):68.

75. Juan NVS, Camilleri M, Jeans JP, Monkhouse A, Chisnall G, Vindrola-Padros C. Redeployment and training of healthcare professionals to intensive care during COVID-19: a systemic review. MedRxiv. 2021:2021.01.21.21250230.

76. Pantoja T, Opiyo N, Lewin S, Paulsen E, Ciapponi A, Wiysonge CS, et al. Implementation strategies for health systems in low-income countries: an overview of systematic reviews. Cochrane Database of Systematic Reviews. 2017(9)n.

77. Ramachandran D, Goswami V, Canny J. Research and reality: using mobile messages to promote maternal health in rural India. Proceedings of the 4th ACM/IEEE International Conference on Information and Communication Technologies and Development; London, United Kingdom: Association for Computing Machinery; 2010. p. Article 35.

78. Moss WJ, Ramakrishnan M, Storms D, Henderson Siegle A, Weiss WM, Lejnev I, Muhe L. Child health in complex emergencies. Bulletin of the World Health Organization. 2006;84:58–64.

79. Bank TW. World Bank Open Data. 2024.

