## Supplemental Document 2 for "Factors impacting on the implementation of clinical management guidelines (CMGs) for high consequence infectious diseases (HCIDs) during outbreaks globally: a systematic review"

**Oxford University Nuffield Department of Medicine: Louise Sigfrid et al**

### Factors influencing the implementation of high consequence infectious disease (HCID) clinical management guidelines (CMGs) during public health emergencies - a rapid qualitative systematic review

*References for the terms in this search strategy:*

1. This search strategy has been informed by the Public Health England search filter for COVID-19: Finding the evidence: Coronavirus. Searching for the evidence [Internet]. Public Health England; 2021 [cited 2021 02 22]. Available from: https://phelibrary.koha-ptfs.co.uk/coronavirusinformation/#Searching
2. The guidelines search filter has been adapted from the CADTH website, the reference for which is as follows: Strings attached: CADTH database search filters [Internet]. Ottawa: CADTH; 2016. [cited 2021 02 22]. Available from: https://www.cadth.ca/resources/finding-evidence/strings-attached-cadths-database-search-filters#guide
3. The implementation terms have been informed by the search strategy for Medline from this reference: Gagliardi, A. R., Marshall, C., Huckson, S., James, R. & Moore, V. 2015. Developing a checklist for guideline implementation planning: review and synthesis of guideline development and implementation advice. *Implementation Science,* 10**,** 19.
4. The qualitative search terms have been informed by the search strategy for Medline from this reference: Search Filters for Various Databases – Ovid Medline [Internet]. The University of Texas School of Public Health; 2020. [cited 2021 02 22]. Available from: https://libguides.sph.uth.tmc.edu/search_filters/ovid_medline_filters

Search strategies run on 09/11/2021 by Eli Harriss, a librarian at the Bodleian Health Care Libraries, University of Oxford. And updated on the 10/11/2021 by Dania Dahmash following the Eli`s strategy.

#### Search Results

| | |
|---|---|
| Ovid Medline | 3,164 |
| Ovid Embase | 2,801 |
| Ovid Global Health | 1,999 |
| Scopus | 2,645 |
| TOTAL | 10,609 |
| Total after deduplication | 9,356 |

#### Search Strategies

Database: Medline (Ovid MEDLINE® Epub Ahead of Print, In-Process & Other Non-Indexed Citations, Ovid MEDLINE® Daily and Ovid MEDLINE®) 1946 to present
Search Strategy:
--------------------------------------------------------------------------------
1 exp clinical pathway/ (7300)
2 exp clinical protocol/ (179667)
3 exp consensus/ (16981)
4 exp consensus development conference/ (12486)
5 exp consensus development conferences as topic/ (2976)
6 critical pathways/ (7300)
7 exp guideline/ (36401)
8 guidelines as topic/ (41717)
9 exp practice guideline/ (29262)
10 practice guidelines as topic/ (126042)
11 (guideline or practice guideline or consensus development conference or consensus development conference, NIH).pt. (46152)
12 (standards or guideline or guidelines).ti,kf,kw. (122209)
13 ((practice or treatment* or clinical) adj guideline*).ab. (45872)
14 (position statement* or policy statement* or practice parameter* or best practice*).ti,ab,kf,kw. (39263)
15 (CPG or CPGs).ti. (6075)
16 consensus*.ti,kf,kw. (30186)
17 ((critical or clinical or practice) adj2 (path or paths or pathway or pathways or protocol*)).ti,ab,kf,kw. (23229)
18 recommendat*.ti,kf,kw. or guideline*.tw. (441428)
19 (care adj2 (standard or path or paths or pathway or pathways or map or maps or plan or plans)).ti,ab,kf,kw. (70116)
20 (algorithm* adj2 (screening or examination or test or tested or testing or assessment* or diagnosis or diagnoses or diagnosed or diagnosing or pharmacotherap* or therap* or treatment* or intervention*)).ti,ab,kf,kw. (20049)
21 1 or 2 or 3 or 4 or 5 or 6 or 7 or 8 or 9 or 10 or 11 or 12 or 13 or 14 or 15 or 16 or 17 or 18 or 19 or 20 (895529)
22 exp Hemorrhagic Fevers, Viral/ (30718)
23 ebolavirus/ or marburgvirus/ (3948)
24 Lassa Fever/ (753)
25 Hemorrhagic Fever Virus, Crimean-Congo/ or Hemorrhagic Fever, Crimean/ (1329)
26 Rift Valley Fever/ (1229)
27 (ebola* or ebov or marburg* or lassa* or CCHF or "crimean-congo*" or "congo virus" or (crimean adj2 (hemorrhagic or haemorrhagic)) or "rift valley*" or RVF).ti,ab. (17834)
28 coronavirus infections/ or COVID-19/ or severe acute respiratory syndrome/ (128393)
29 ("middle east* respiratory syndr*" or MERS-CoV or "novel CoV*" or "novel betacoronavirus" or "novel coronavirus*" or ("middle east" adj3 (coronavirus* or cov or betacoronavirus*)) or (MERS adj3 (coronavirus* or cov or betacoronavirus*)) or "mers-coronavirus" or "mers cov" or merscov).tw. (14235)
30 ("wuhan flu" or 2019-nCoV).tw. (1395)
31 (h1n1 or h5n1).tw. (24011)
32 "zoonotic influenza*".tw. (132)
33 Influenza A Virus, H1N1 Subtype/ (16441)
34 Influenza A Virus, H5N1 Subtype/ (6253)
35 exp Henipavirus/ (817)
36 Henipavirus Infections/ (573)
37 (nipah or hendra).tw. (1365)
38 Monkeypox virus/ or Monkeypox/ (497)
39 (monkeypox or "monkey pox").tw. (805)
40 Chikungunya virus/ or Chikungunya Fever/ (3847)
41 Chikungunya.tw. (6027)
42 "Severe Fever with Thrombocyto* Syndrome".tw. (716)

43 SFTS.tw. (1069)
44 Plague/ (5403)
45 plague.tw. (9625)
46 ("black death" or "pathogen x").tw. (337)
47 Dengue Virus/ or exp Dengue/ or Dengue.tw. (25706)
48 communicable diseases, emerging/ or communicable diseases, imported/ or ("infectious diseases" or "dangerous pathogen*").ti,ab. (66980)
49 exp Influenza A virus/ (46577)
50 Influenza, Human/ (53364)
51 (H1N1 or H2N2 or H3N2 or H5N1 or H5N8 or H7N3 or H7N9 or ((influenza or flu) adj3 (pandemic* or novel or new or outbreak*))).tw,kf. (37129)
52 exp Coronavirus/ (105839)
53 ((corona* or corono*) adj1 (virus* or viral* or virinae*)).tw. (2600)
54 (coronavirus* or coronovirus* or coronavirinae* or CoV or covid*).tw. (205057)
55 ("2019-nCoV" or 2019nCoV or nCoV2019 or "nCoV-2019" or nCoV19 or "nCoV-19" or "COVID-19" or COVID19 or "CORVID-19" or CORVID19 or "WN-CoV" or WNCoV or "HCoV-19" or HCoV19 or "HCoV-2019" or HCoV2019 or "2019 novel*" or Ncov or "n-cov" or "SARS-CoV-2" or "SARSCoV-2" or "SARSCoV2" or "SARS-CoV2" or SARSCov19 or "SARS-Cov19" or "SARSCov-19" or "SARS-Cov-19" or Ncovor or Ncorona* or Ncorono* or NcovWuhan* or NcovHubei* or NcovChina* or NcovChinese* or SARS2 or "SARS-2" or SARScoronavirus2 or "SARS-coronavirus-2" or "SARScoronavirus 2" or "SARS coronavirus2" or SARScoronovirus2 or "SARS-coronovirus-2" or "SARScoronovirus 2" or "SARS coronovirus2" or SARS).tw. (192047)
56 (respiratory* adj2 (symptom* or disease* or illness* or condition*) adj10 (Wuhan* or Hubei* or China* or Chinese* or Huanan*)).tw. (666)
57 (pneumonia* adj10 (Wuhan* or Hubei* or China* or Chinese* or Huanan*)).tw. (1998)
58 disease outbreaks/ or disease hotspot/ or epidemics/ or pandemics/ or space-time clustering/ or (outbreak* or pandemic* or epidemic* or endemic or (("public health" or "global health") adj4 (emergency or emergencies))).tw. (448842)
59 "severe acute respiratory syndrome*".tw. (25833)
60 MERS.tw. (6717)
61 SARS Virus/ (3983)
62 SARS.tw. (66909)
63 22 or 23 or 24 or 25 or 26 or 27 or 28 or 29 or 30 or 31 or 32 or 33 or 34 or 35 or 36 or 37 or 38 or 39 or 40 or 41 or 42 or 43 or 44 or 45 or 46 or 47 or 48 or 49 or 50 or 51 or 52 or 53 or 54 or 55 or 56 or 57 or 58 or 59 or 60 or 61 or 62 (690485)
64 Guideline Adherence/ (34264)
65 (implement* or adher* or diffus* or utilise* or adapt* or develop* or deploy* or "use" or using or complian* or utility or utilisation or utilization or disseminat* or translat* or adopt* or aware* or uptake or up?take or influenc* or follow or comply or complying or barrier* or facilitat*).tw. (14222548)
66 64 or 65 (14230793)
67 exp "Attitude of Health Personnel"/ (165703)
68 exp Health Personnel/ (563859)
69 exp Patient Care Team/ (71342)
70 (staff or "healthcare workforce" or physician* or clinician* or stakeholders or "healthcare professional*").ti,ab. (857988)
71 (doctor* or clinician* or "family practition*" or "general practition*" or nurs* or obstetrician* or physician* or neurologist* or "health profession*" or (health?care adj2 (profession* or worker* or provider* or staff or personnel)) or "nursing staff" or "medical assist*" or "public health practition*" or consultant* or "care practition*" or "medical practition*" or p?ediatric*).tw. (1708229)
72 67 or 68 or 69 or 70 or 71 (2187900)
73 exp qualitative research/ (69794)
74 interview/ (29973)
75 focus groups/ or interviews as topic/ or exp "surveys and questionnaires"/ (1206843)
76 (qualitative* or survey* or "focus group*" or interview*).tw. (1278747)
77 (phenomenological or experienc* or perception* or perceiv* or "grounded theory").tw. (1631328)
78 perspective*.tw. (371418)
79 ((("semi-structured" or semistructured or unstructured or informal or "in-depth" or indepth or "face-to-face" or structured or guide) adj3 (interview* or discussion* or questionnaire*)) or (focus group* or qualitative or ethnograph* or fieldwork or "field work" or "key informant")).ti,ab. or interviews as topic/ or narration/ or "pre-post".tw. (446896)

80 73 or 74 or 75 or 76 or 77 or 78 or 79 (3563864)
81 21 and 63 and 66 and 72 and 80 (3164)

**Database: Embase 1974 to present**
Search Strategy:
--------------------------------------------------------------------------------
1 ((guideline* or protocol* or path or paths or pathway* or consensus* or standards or position statement* or policy statement* or practice parameter* or best practice* or CPG or CPGs or recommendat* or standard or map or maps or plan or plans or algorithm*) adj3 (implement* or adher* or diffus* or utilise* or adapt* or develop* or deploy* or "use" or using or complian* or utility or utilisation or utilization or disseminat* or translat* or adopt* or aware* or uptake or up?take or influenc* or follow or comply or complying or barrier* or facilitat*)).tw. (644840)
2 Ebola hemorrhagic fever/ (6975)
3 exp ebolavirus/ (4062)
4 Marburgvirus/ or Marburg hemorrhagic fever/ or Marburg virus/ (1453)
5 Lassa virus/ or Lassa fever/ (1866)
6 Crimean-Congo hemorrhagic fever virus/ or Crimean Congo hemorrhagic fever/ (1666)
7 Rift Valley fever virus/ or Rift Valley fever bunyavirus/ or Rift Valley fever/ (2113)
8 (ebola* or ebov or marburg* or lassa* or CCHF or "crimean-congo*" or "congo virus" or (crimean adj2 (hemorrhagic or haemorrhagic)) or "rift valley*" or RVF).ti,ab. (21115)
9 exp coronavirinae/ (69363)
10 exp Coronavirus infection/ (181493)
11 Middle East respiratory syndrome coronavirus/ or Middle East respiratory syndrome/ (5744)
12 ("middle east* respiratory syndr*" or MERS-CoV or "novel CoV*" or "novel betacoronavirus" or "novel coronavirus*" or ("middle east" adj3 (coronavirus* or cov or betacoronavirus*)) or (MERS adj3 (coronavirus* or cov or betacoronavirus*)) or "mers-coronavirus" or "mers cov" or merscov).tw. (14589)
13 ("wuhan flu" or 2019-nCoV).tw. (1429)
14 (h1n1 or h5n1).tw. (29677)
15 "zoonotic influenza*".tw. (154)
16 exp "influenza a virus (h1n1)"/ (5342)
17 "influenza a virus (h5n1)"/ (1585)
18 Henipavirus infection/ or Henipavirus/ (393)
19 (nipah or hendra).tw. (1541)
20 monkeypox/ or Monkeypox virus/ (861)
21 (monkeypox or "monkey pox").tw. (896)
22 Chikungunya virus/ or chikungunya/ (6130)
23 Chikungunya.tw. (7404)
24 severe fever with thrombocytopenia syndrome/ (538)
25 "Severe Fever with Thrombocyto* Syndrome".tw. (771)
26 SFTS.tw. (1272)
27 plague/ (6342)
28 plague.tw. (8470)
29 ("black death" or "pathogen x").tw. (333)
30 dengue hemorrhagic fever/ or dengue/ or exp Dengue virus/ (31200)
31 Dengue.tw. (30770)
32 ("infectious diseases" or "dangerous pathogen*").ti,ab. (89205)
33 exp influenza a virus/ (15966)
34 (H1N1 or H2N2 or H3N2 or H5N1 or H5N8 or H7N3 or H7N9 or ((influenza or flu) adj3 (pandemic* or novel or new or outbreak*))).tw. (43988)
35 ((corona* or corono*) adj1 (virus* or viral* or virinae*)).tw. (3050)
36 (coronavirus* or coronovirus* or coronavirinae* or CoV or covid*).tw. (211223)
37 ("2019-nCoV" or 2019nCoV or nCoV2019 or "nCoV-2019" or nCoV19 or "nCoV-19" or "COVID-19" or COVID19 or "CORVID-19" or CORVID19 or "WN-CoV" or WNCoV or "HCoV-19" or HCoV19 or "HCoV-2019" or HCoV2019 or "2019 novel*" or Ncov or "n-cov" or "SARS-CoV-2" or "SARSCoV-2" or "SARSCoV2" or "SARS-CoV2" or SARSCov19 or "SARS-Cov19" or "SARSCov-19" or "SARS-Cov-19" or Ncovor or Ncorona* or Ncorono* or NcovWuhan* or NcovHubei* or NcovChina* or NcovChinese* or SARS2 or "SARS-2" or SARScoronavirus2 or "SARS-coronavirus-2" or

"SARScoronavirus 2" or "SARS coronavirus2" or SARScoronovirus2 or "SARS-coronovirus-2" or "SARScoronovirus 2" or "SARS coronovirus2" or SARS).tw. (195180)
38 (respiratory* adj2 (symptom* or disease* or illness* or condition*) adj10 (Wuhan* or Hubei* or China* or Chinese* or Huanan*)).tw. (801)
39 (pneumonia* adj10 (Wuhan* or Hubei* or China* or Chinese* or Huanan*)).tw. (2291)
40 epidemic/ (116941)
41 pandemic/ (94322)
42 (outbreak* or pandemic* or epidemic* or endemic or (("public health" or "global health") adj4 (emergency or emergencies))).tw. (433649)
43 "severe acute respiratory syndrome*".tw. (25310)
44 MERS.tw. (7270)
45 SARS.tw. (68683)
46 2 or 3 or 4 or 5 or 6 or 7 or 8 or 9 or 10 or 11 or 12 or 13 or 14 or 15 or 16 or 17 or 18 or 19 or 20 or 21 or 22 or 23 or 24 or 25 or 26 or 27 or 28 or 29 or 30 or 31 or 32 or 33 or 34 or 35 or 36 or 37 or 38 or 39 or 40 or 41 or 42 or 43 or 44 or 45 (747977)
47 exp health care personnel/ (1722002)
48 (staff or "healthcare workforce" or physician* or clinician* or stakeholders or "healthcare professional*").ti,ab. (1204922)
49 (doctor* or clinician* or "family practition*" or "general practition*" or nurs* or obstetrician* or physician* or neurologist* or "health profession*" or (health?care adj2 (profession* or worker* or provider* or staff or personnel)) or "nursing staff" or "medical assist*" or "public health practition*" or consultant* or "care practition*" or "medical practition*" or p?ediatric*).tw. (2315153)
50 47 or 48 or 49 (3382207)
51 exp qualitative research/ or qualitative analysis/ (161060)
52 exp interview/ or exp questionnaire/ (1062906)
53 ethnography/ (3149)
54 (qualitative* or survey* or "focus group*" or interview*).tw. (1612840)
55 (phenomenological or experienc* or perception* or perceiv* or "grounded theory").tw. (2192722)
56 perspective*.tw. (446679)
57 ((("semi-structured" or semistructured or unstructured or informal or "in-depth" or indepth or "face-to-face" or structured or guide) adj3 (interview* or discussion* or questionnaire*)) or (focus group* or qualitative or ethnograph* or fieldwork or "field work" or "key informant" or pre-post*)).ti,ab. (519296)
58 51 or 52 or 53 or 54 or 55 or 56 or 57 (4162786)
59 1 and 46 and 50 and 58 (2801)

**Database: Global Health <1973 to 2021 Week 45>**
Search Strategy:
--------------------------------------------------------------------------------
1 consensus/ (152)
2 guidelines/ (56501)
3 (standards or guideline or guidelines).ti,ab. (119077)
4 (position statement* or policy statement* or practice parameter* or best practice*).ti,ab. (5970)
5 (CPG or CPGs).ti. (537)
6 consensus*.ti,ab. (19804)
7 ((critical or clinical or practice) adj2 (path or paths or pathway or pathways or protocol*)).ti,ab. (1917)
8 recommendat*.ti,ab. (70799)
9 (care adj2 (standard or path or paths or pathway or pathways or map or maps or plan or plans)).ti,ab. (6927)
10 (algorithm* adj2 (screening or examination or test or tested or testing or assessment* or diagnosis or diagnoses or diagnosed or diagnosing or pharmacotherap* or therap* or treatment* or intervention*)).ti,ab. (2175)
11 1 or 2 or 3 or 4 or 5 or 6 or 7 or 8 or 9 or 10 (192119)
12 exp viral haemorrhagic fevers/ (7304)
13 exp ebolavirus/ or exp marburgvirus/ or exp lassa virus/ (5880)
14 exp crimean-congo haemorrhagic fever virus/ (1741)
15 exp rift valley fever virus/ (1886)

16 (ebola* or ebov or marburg* or lassa* or CCHF or "crimean-congo*" or "congo virus" or (crimean adj2 (hemorrhagic or haemorrhagic)) or "rift valley*" or RVF).ti,ab. (10301)
17 severe acute respiratory syndrome.sh. or Severe acute respiratory syndrome coronavirus.od. (7716)
18 ("middle east* respiratory syndr*" or MERS-CoV or "novel CoV*" or "novel betacoronavirus" or "novel coronavirus*" or ("middle east" adj3 (coronavirus* or cov or betacoronavirus*)) or (MERS adj3 (coronavirus* or cov or betacoronavirus*)) or "mers-coronavirus" or "mers cov" or merscov).tw. (6210)
19 ("wuhan flu" or 2019-nCoV).tw. (1325)
20 (h1n1 or h5n1).tw. (14116)
21 "zoonotic influenza*".tw. (130)
22 exp influenza a virus/ (20909)
23 exp henipavirus/ (1154)
24 (nipah or hendra).tw. (1165)
25 exp monkeypox virus/ (389)
26 (monkeypox or "monkey pox").tw. (490)
27 exp Chikungunya virus/ (4509)
28 Chikungunya.tw. (5303)
29 "Severe Fever with Thrombocyto* Syndrome".tw. (576)
30 SFTS.tw. (533)
31 plague/ (3352)
32 plague.tw. (7693)
33 ("black death" or "pathogen x").tw. (116)
34 exp dengue virus/ or Dengue.tw. (22895)
35 emerging infectious diseases/ or ("infectious diseases" or "dangerous pathogen*").ti,ab. (35565)
36 ((corona* or corono*) adj1 (virus* or viral* or virinae*)).tw. (1152)
37 (coronavirus* or coronovirus* or coronavirinae* or CoV or covid*).tw. (59248)
38 ("2019-nCoV" or 2019nCoV or nCoV2019 or "nCoV-2019" or nCoV19 or "nCoV-19" or "COVID-19" or COVID19 or "CORVID-19" or CORVID19 or "WN-CoV" or WNCoV or "HCoV-19" or HCoV19 or "HCoV-2019" or HCoV2019 or "2019 novel*" or Ncov or "n-cov" or "SARS-CoV-2" or "SARSCoV-2" or "SARSCoV2" or "SARS-CoV2" or SARSCov19 or "SARS-Cov19" or "SARSCov-19" or "SARS-Cov-19" or Ncovor or Ncorona* or Ncorono* or NcovWuhan* or NcovHubei* or NcovChina* or NcovChinese* or SARS2 or "SARS-2" or SARScoronavirus2 or "SARS-coronavirus-2" or "SARScoronavirus 2" or "SARS coronavirus2" or SARScoronovirus2 or "SARS-coronovirus-2" or "SARScoronovirus 2" or "SARS coronovirus2" or SARS).tw. (55013)
39 (respiratory* adj2 (symptom* or disease* or illness* or condition*) adj10 (Wuhan* or Hubei* or China* or Chinese* or Huanan*)).tw. (1028)
40 (pneumonia* adj10 (Wuhan* or Hubei* or China* or Chinese* or Huanan*)).tw. (1581)
41 outbreaks/ or exp epidemics/ or (outbreak* or pandemic* or epidemic* or endemic or (("public health" or "global health") adj4 (emergency or emergencies))).tw. (211102)
42 "severe acute respiratory syndrome*".tw. (51955)
43 MERS.tw. (2756)
44 Middle East respiratory syndrome Coronavirus.od. (1902)
45 SARS.tw. (26727)
46 12 or 13 or 14 or 15 or 16 or 17 or 18 or 19 or 20 or 21 or 22 or 23 or 24 or 25 or 26 or 27 or 28 or 29 or 30 or 31 or 32 or 33 or 34 or 35 or 36 or 37 or 38 or 39 or 40 or 41 or 42 or 43 or 44 or 45 (296241)
47 (implement* or adher* or diffus* or utilise* or adapt* or develop* or deploy* or "use" or using or complian* or utility or utilisation or utilization or disseminat* or translat* or adopt* or aware* or uptake or up?take or influenc* or follow or comply or complying or barrier* or facilitat*).tw. (2927726)
48 exp health care workers/ or (staff or "healthcare workforce" or physician* or clinician* or stakeholders or "healthcare professional*").ti,ab. (167975)
49 (doctor* or clinician* or "family practition*" or "general practition*" or nurs* or obstetrician* or physician* or neurologist* or "health profession*" or (health?care adj2 (profession* or worker* or provider* or staff or personnel)) or "nursing staff" or "medical assist*" or "public health practition*" or consultant* or "care practition*" or "medical practition*" or p?ediatric*).tw. (207519)
50 48 or 49 (265554)
51 qualitative analysis/ or qualitative techniques/ (4318)
52 exp interviews/ or exp questionnaires/ or exp surveys/ or ethnography/ (142857)
53 (qualitative* or survey* or "focus group*" or interview*).tw. (408892)
54 (phenomenological or experienc* or perception* or perceiv* or "grounded theory").tw. (221796)

55 perspective*.tw. (56474)
56 ((("semi-structured" or semistructured or unstructured or informal or "in-depth" or indepth or "face-to-face" or structured or guide) adj3 (interview* or discussion* or questionnaire*)) or (focus group* or qualitative or ethnograph* or fieldwork or "field work" or "key informant")).ti,ab. or interviews as topic/ or narration/ or "pre-post".tw. (105225)
57 51 or 52 or 53 or 54 or 55 or 56 (615378)
58 11 and 46 and 47 and 50 and 57 (1999)

**Scopus**

( ( TITLE-ABS-KEY ( ( ( guideline* OR protocol* OR path OR paths OR pathway* OR consensus* ) W/3 ( implement* OR adher* OR diffus* OR utilise* OR adapt* OR develop* OR deploy* OR use OR using OR complian* OR utility OR utilisation OR utilization OR disseminat* OR translat* OR adopt* OR aware* OR uptake OR up-take OR influenc* OR follow OR comply OR complying OR barrier* OR facilitat* ) ) ) ) OR ( TITLE-ABS-KEY ( standards W/3 ( implement* OR adher* OR diffus* OR utilise* OR adapt* OR develop* OR deploy* OR use OR using OR complian* OR utility OR utilisation OR utilization OR disseminat* OR translat* OR adopt* OR aware* OR uptake OR up-take OR influenc* OR follow OR comply OR complying OR barrier* OR facilitat* ) ) ) OR ( TITLE-ABS-KEY ( ( "position statement*" OR "policy statement*" OR "practice parameter*" OR "best practice*" ) W/3 ( implement* OR adher* OR diffus* OR utilise* OR adapt* OR develop* OR deploy* OR use OR using OR complian* OR utility OR utilisation OR utilization OR disseminat* OR translat* OR adopt* OR aware* OR uptake OR up-take OR influenc* OR follow OR comply OR complying OR barrier* OR facilitat* ) ) ) OR ( TITLE-ABS-KEY ( ( cpg OR cpgs OR recommendat* OR standard OR map OR maps OR plan OR plans OR algorithm* ) W/3 ( implement* OR adher* OR diffus* OR utilise* OR adapt* OR develop* OR deploy* OR use OR using OR complian* OR utility OR utilisation OR utilization OR disseminat* OR translat* OR adopt* OR aware* OR uptake OR up-take OR influenc* OR follow OR comply OR complying OR barrier* OR facilitat* ) ) ) ) AND ( ( TITLE-ABS-KEY ( ebola* OR ebov OR marburg* OR lassa* OR cchf OR "crimean-congo*" OR "congo virus" OR ( crimean W/2 ( hemorrhagic OR haemorrhagic ) ) OR "rift valley*" OR rvf ) ) OR ( TITLE-ABS-KEY ( ( "middle east* respiratory syndr*" OR mers-cov OR "novel CoV*" OR "novel betacoronavirus" OR "novel coronavirus*" OR ( "middle east" W/3 ( coronavirus* OR cov OR betacoronavirus* ) ) OR ( mers W/3 ( coronavirus* OR cov OR betacoronavirus* ) ) OR "mers-coronavirus" OR "mers cov" OR merscov ) ) ) OR ( TITLE-ABS-KEY ( "wuhan flu" OR 2019-ncov OR h1n1 OR h5n1 OR "zoonotic influenza*" OR ( nipah OR hendra ) OR ( monkeypox OR "monkey pox" ) OR chikungunya OR "Severe Fever with Thrombocyto* Syndrome" OR sfts ) ) OR ( TITLE-ABS-KEY ( plague OR "black death" OR "pathogen x" OR dengue OR "infectious diseases" OR "dangerous pathogen*" OR h1n1 OR h2n2 OR h3n2 OR h5n1 OR h5n8 OR h7n3 OR h7n9 OR ( ( influenza OR flu ) W/3 ( pandemic* OR novel OR new OR outbreak* ) ) ) ) ) OR ( TITLE-ABS-KEY ( ( ( corona* OR corono* ) W/1 ( virus* OR viral* OR virinae* ) ) ) ) OR ( TITLE-ABS-KEY ( ( coronavirus* OR coronovirus* OR coronavirinae* OR cov OR covid* OR "2019-nCoV" OR 2019ncov OR ncov2019 OR "nCoV-2019" OR ncov19 OR "nCoV-19" OR "COVID-19" OR covid19 OR "CORVID-19" OR corvid19 OR "WN-CoV" OR wncov OR "HCoV-19" OR hcov19 OR "HCoV-2019" OR hcov2019 OR "2019 novel*" OR ncov OR "n-cov" OR "SARS-CoV-2" OR "SARSCoV-2" OR "SARSCoV2" OR "SARS-CoV2" OR sarscov19 OR "SARS-Cov19" OR "SARSCov-19" OR "SARS-Cov-19" OR ncovor OR ncorona* OR ncorono* OR ncovwuhan* OR ncovhubei* OR ncovchina* OR ncovchinese* OR sars2 OR "SARS-2" OR sarscoronavirus2 OR "SARS-coronavirus-2" OR "SARScoronavirus 2" OR "SARS coronavirus2" OR sarscoronovirus2 OR "SARS-coronovirus-2" OR "SARScoronovirus 2" OR "SARS coronovirus2" OR sars ) ) ) OR ( TITLE-ABS-KEY ( ( respiratory* W/2 ( symptom* OR disease* OR illness* OR condition* ) W/10 ( wuhan* OR hubei* OR china* OR chinese* OR huanan* ) ) ) ) OR ( TITLE-ABS-KEY ( ( pneumonia* W/10 ( wuhan* OR hubei* OR china* OR chinese* OR huanan* ) ) ) ) OR ( TITLE-ABS-KEY ( ( outbreak* OR pandemic* OR epidemic* OR endemic OR ( ( "public health" OR "global health" ) W/4 ( emergency OR emergencies ) ) ) ) ) OR ( TITLE-ABS-KEY ( "severe acute respiratory syndrome*" OR sars OR mers ) ) ) AND ( ( TITLE-ABS-KEY ( staff OR "healthcare workforce" OR physician* OR clinician* OR stakeholders OR "healthcare professional*" ) ) OR ( TITLE-ABS-KEY ( doctor* OR clinician* OR "family practition*" OR "general practition*" OR nurs* OR obstetrician* OR physician* OR neurologist* OR "health profession*" ) ) OR ( TITLE-ABS-KEY ( ( ( healthcare OR "health care" ) W/2 ( profession* OR worker* OR provider* OR staff OR personnel ) ) ) ) OR ( TITLE-ABS-KEY ( "nursing staff" OR "medical assist*" OR "public health practition*" OR consultant* OR "care practition*" OR "medical practition*" OR pediatrician* OR paediatrician* ) ) ) AND ( ( TITLE-ABS-KEY ( qualitative* OR survey* OR "focus group*" OR interview* ) ) OR ( TITLE-ABS-KEY ( phenomenological OR experienc* OR perception* OR perceiv* OR "grounded theory" ) ) OR ( TITLE-ABS-KEY ( perspective* ) ) OR ( TITLE-ABS-KEY ( ( ( "semi-structured" OR semistructured OR unstructured OR informal OR "in-depth" OR indepth OR "face-to-face" OR structured OR guide ) W/3 ( interview* OR discussion* OR questionnaire* ) ) ) ) OR ( TITLE-ABS-KEY ( "focus group*" OR qualitative OR ethnograph* OR fieldwork OR "field work" OR "key informant" OR pre-post* ) ) )
