## Supplemental Document 3 for "Factors impacting on the implementation of clinical management guidelines (CMGs) for high consequence infectious diseases (HCIDs) during outbreaks globally: a systematic review"

**Appendix 1: Characteristics of included studies (N=28) grouped by HCID type**

| Study | Study setting, Income classification (79) | Study design | Participants | HCID | Key themes Identified | Risk of bias |
|---|---|---|---|---|---|---|
| Alanezi, F. et al. (2021) (37) | Saudi Arabia (HIC) | Qualitative | Nurses (28.4%), senior physicians (23.5%), assistant physicians (12.7%), consultants or chief physicians or medical students (10.8%), pharmacists or administrators (8.8%), others (n=102). | COVID-19 | -Logistics organisational support | High |
| Chuang, E.et al. (2020) (27) | USA (HIC) | Qualitative | Palliative care focus group: Social workers, physicians (n=14).<br>Critical care focus group: physician fellows, junior attending physicians, senior attending physicians, and intensive care unit leadership (n=25).<br>Nursing focus group (n=4).<br>Respiratory therapist (n=20).<br>Interviews: one academic hospital medical director, directors of emergency medicine, and one respiratory therapist. | COVID-19 | -Guidelines characteristics, development, and dissemination<br>-Logistics organisational support<br>-Realities of implementing guidelines | High |
| Driver, J. A. et al. (2020) (45) | USA (HIC) | Quantitative | Veterans received outpatient monitoring (n=120) | COVID-19 | -Logistics organisational support | Moderate |
| Francisco, M. A. et al. (2020) (40) | USA (HIC) | Quantitative | Nurses (n=78) | COVID-19 | -Logistics organisational support<br>-Realities of implementing guidelines | Moderate |
| Gonzalez, A. M. et al. (2020) (32) | Spain (HIC) | Qualitative | Clinical cases (n=5) | COVID-19 | -Guidelines characteristics, development, and dissemination<br>-Logistics organisational support<br>-Realities of implementing guidelines | Moderate |
| Kurotschka, P. K. et al. (2020) (30) | Italy (HIC) | Qualitative | General practitioners (n=149) | COVID-19 | -Guidelines characteristics, development, and dissemination | High |
| Alqahtani, J. S. et al. (2020) (43) | Based in UK ** (HIC) | Quantitative | Critical care practitioners (n=502) | COVID-19 | -Logistics organisational support | Moderate |

| | | | | | | |
|---|---|---|---|---|---|---|
| Ismail, M. et al. (2021) (44) | Qatar (HIC) | Quantitative | Primary care physicians (n=294) | COVID-19 | -Guidelines characteristics, development, and dissemination | High |
| Balkrishnan, P et al. (2019) (38) | India (LMIC) | Quantitative | Patients (pre-intervention phase of six months having 84 patients and post-intervention phase of six months having 50 patients) (n= 134). | Dengue | -Logistics organisational support | Moderate |
| Handel, A. S. et al. (2016) (41) | Ecuador (Upper-Middle Income) | Quantitative | Healthcare providers (n=76) medical doctors (82 %)<br>Nurses (14 %). | Dengue | -Logistics organisational support | High |
| Kularatne, S. A. et al. (2003) (42) | Sri-Lanka (LMIC) | Quantitative | Consultant physicians(n=35)<br>Consultant paediatricians (n=15). | Dengue | -Guidelines characteristics, development, and dissemination<br>-Realities of implementing guidelines | Low |
| Wharton-Smith, A.et al. (2014) (34) | Malaysia (Upper-Middle Income) | Qualitative | Medical officers. Six FGDs: (n= 54).<br>Interviews (n=14). | Dengue | -Guidelines characteristics, development, and dissemination<br>-Logistics organisational support<br>-Realities of implementing guidelines | High |
| Barniol, J. et al. (2011) (46) | 18 countries in four WHO regions | Mixed method | Participants (n=1288)<br>Clinicians, nurses, communicable disease control staff, epidemiologists, management from health care facilities.<br>Chart review total (n=3248). | Dengue | -Logistics organisational support<br>-Realities of implementing guidelines | High |
| Raffetin, A. et al. (2018) (49) | France (HIC) | Mixed method | Family physician (n=33) | EVD | -Guidelines characteristics, development, and dissemination | Moderate |
| Christine Loignon et al. (2018) (36) | West Africa | Qualitative | Stakeholders (decision-makers, physicians, nurses). (n=29). | EVD | Logistics organisational support | High |
| Battista, M. C.et al. (2019) (39) | West Africa | Quantitative | Physicians (n= 24, (83%)), nurses (n= 3, 10%), and project management and coordination (n=2, 7%). Total sample: n= 29. | EVD | -Guidelines characteristics, development, and dissemination<br>-Logistics organisational support | Moderate |
| Broom, J.et al. (2016) (26) | Australia (HIC) | Qualitative | Consultants (n=8), and nurses (n=13).<br>Total sample: n= 21. | EVD | -Guidelines characteristics, development, and dissemination | High |
| Erland, E. et al. (2017) (28) | West Africa | Qualitative | Midwives (n=11) | EVD | -Guidelines characteristics, development, and dissemination.<br>-Realities of implementing guidelines | High |

| | | | | | | |
|---|---|---|---|---|---|---|
| Lam, S. et al. (2014) (31) | Hong Kong (HIC) | Qualitative | Emergency nurses (n=12) | Emerging infectiou s diseases | -Realities of implementing guidelines | High |
| Nhan, C.et al. (2012) (48) | Canada (HIC) | Mixed method | 39% (68/ 173) and 41% (61/147) of AMMIQ and AMSSCQ members, respectively, (n =129). | Influenza (H1N1) | -Guidelines characteristics, development, and dissemination<br>-Logistics organisational support<br>-Realities of implementing guidelines | Moderate |
| Filice, C. E.et al. (2013) (29) | USA (HIC) | Qualitative | Medical directors (n=15) and designated other EPs in leadership roles in their departments (n=5). | Influenza (H1N1) | -Guidelines characteristics, development, and dissemination<br>-Logistics organisational support<br>-Realities of implementing guidelines | High |
| Ahankari, A. S et al. (2017) (25) | India (LMIC) | Qualitative | General physicians (GPs), specialists in allopathy (n = 16), Ayurveda (n = 3) homeopathy (n = 1). Total practitioners (n=20). | Influenza | -Guidelines characteristics, development, and dissemination | High |
| Haq, Z et al. (2016) (51) | Mediterranean Region | Opinion piece | Not applicable | Influenza | -Guidelines characteristics, development, and dissemination<br>-Logistics organisational support<br>-Realities of implementing guidelines | * |
| Gesser-Edelsburg et al. (2014) (35) | Israel (HIC) | Qualitative | Stakeholders (hospital physicians (n=14), hospital nurse (n=21), community physicians (n= 10), community nurse (n=4). Total sample (n=70). | Influenza | -Logistics organisational support<br>-Realities of implementing guidelines | High |
| Phipps, E.et al. (2019) (33) | England (HIC) | Qualitative | Outbreaks (n=109). | Influenza | -Guidelines characteristics, development, and dissemination | High |
| Bandara, T. et al. (2020) (50) | Canada (HIC) | Opinion piece | Not applicable | Influenza | -Guidelines characteristics, development, and dissemination<br>-Logistics organisational support<br>-Realities of implementing guidelines | * |
| Omrani, A. S.et al. (2015) (52) | Saudi Arabia (HIC) | Opinion piece | Not applicable | MERS-CoV | -Realities of implementing guidelines | * |
