## Supplemental Document 4 for "Factors impacting on the implementation of clinical management guidelines (CMGs) for high consequence infectious diseases (HCIDs) during outbreaks globally: a systematic review"

**Appendix 2-a: Quality appraisal of included studies using the MMAT Qualitative tool**

| Study | 1.1. Is the qualitative approach appropriate to answer the research question? | 1.2. Are the qualitative data collection methods adequate to address the research question? | 1.3. Are the findings adequately derived from the data? | 1.4. Is the interpretation of results sufficiently substantiated by data? | 1.5. Is there coherence between qualitative data sources collection, analysis and interpretation? |
|---|---|---|---|---|---|
| Ahankari et al.(25) | Yes | Yes | Yes | Yes | Yes |
| Broom et al.(26) | Yes | Yes | Yes | Yes | Yes |
| Chuang et al.(27) | Yes | Yes | Yes | Yes | Yes |
| Erland et al.(28) | Yes | Yes | Yes | Yes | Yes |
| Filice et al.(29) | Yes | Yes | Yes | Yes | Yes |
| Kurotschka et al.(30) | Yes | Yes | Yes | Yes | Yes |
| Lam et al.(31) | Yes | Yes | Yes | Yes | Yes |
| Gonzalez et al.(32) | Yes | Can't tell | Yes | Yes | Yes |
| Phipps et al.(33) | Yes | Yes | Yes | Yes | Yes |
| Wharton-Smith et al.(34) | Yes | Yes | Yes | Yes | Yes |
| Gesser-Edelsburg et al.(35) | Yes | Yes | Yes | Yes | Yes |
| Loignon et al.(36) | Yes | Yes | Yes | Yes | Yes |

**Appendix 2-b: Quality appraisal of included studies using the MMAT quantitative tools**

| Study | Quantitative studies | | | | |
|---|---|---|---|---|---|
| | 2.1. Is the sampling strategy relevant to address the research question? | 2.2. Is the sample representative of the target population? | 2.3. Are the measurements appropriate? | 2.4. Is the risk of nonresponse bias low? | 2.5. Is the statistical analysis appropriate to answer the research question? |
| Alanezi F et al.(37) | Yes | Yes | Yes | Yes | Yes |
| Balkrishnan, P et al.(38) | Yes | Yes | Yes | Can’t tell | Yes |
| Battista, M. C.et al.(39) | Yes | Yes | Yes | Can’t tell | Yes |
| Francisco M. A et al.(40) | Yes | Yes | Yes | No | Yes |
| Handel, A. S. et al.(41) | Yes | Yes | Yes | Yes | Yes |
| Kularatne, S. A. et al.(42) | No | No | No | Yes | No |

| | | | | | |
|---|---|---|---|---|---|
| **Alqahtani, J. S.et al.(43)** | Yes | No | Yes | No | Yes |
| **Ismail, M. et al.(44)** | Yes | Yes | Yes | Yes | Yes |
| | **Quantitative non-randomized** | | | | |
| | **3.1. Are the participants representative of the target population?** | **3.2. Are measurements appropriate regarding both the outcome intervention or exposure?** | **3.3. Are there complete outcome data?** | **3.4. Are the confounders accounted for in the design and analysis?** | **3.5. During the study period is the intervention administered or exposure occurred as intended?** |
| **Driver, J. A. et al.(45)** | Yes | Yes | Can't tell | Can't tell | Yes |

### Appendix 2-c: Quality appraisal of included studies using the MMAT mixed method tool

| Study | 4.1. Is there an adequate rationale for using a mixed methods design to address the research question? | 4.2. Are the different components of the study effectively integrated to answer the research question? | 4.3. Are the outputs of the integration of qualitative and quantitative components adequately interpreted? | 4.4. Are divergences and inconsistencies between quantitative and qualitative results adequately addressed? | 4.5. Do the different components of the study adhere to the quality criteria of each tradition of the methods involved? |
|---|---|---|---|---|---|
| **Barniol, J. et al.(46)** | Yes | Yes | Yes | Yes | Yes |
| **Fryk, J. J. et al.(47)** | Yes | Yes | Yes | Yes | Yes |
| **Nhan, C. et al.(48)** | Yes | Yes | Yes | No | No |
| **Raffetin, A. et al.(49)** | Yes | Yes | Yes | No | Yes |

### Appendix 2-d: Quality appraisal of included studies using the JBI critical appraisal checklist for text and opinion papers

| Study | Is the source of the opinion clearly identified? | Does the source of opinion have standing in the field of expertise? | Are the interests of the relevant population the central focus of the opinion? | Is the stated position the result of an analytical process and is there logic in the opinion expressed? | Is there reference to the extant literature? | Is any incongruence with the literature sources logically defended? |
|---|---|---|---|---|---|---|
| **Bandara, T. et al.(50)** | Yes | Yes | Yes | Yes | Yes | Yes |
| **Haq, Z. et al.(51)** | Yes | Yes | Yes | Yes | Yes | Yes |
| **Omrani, A. S. et al.(52)** | Yes | Yes | Yes | Yes | Yes | Yes |
